# Seroprevalence, Environmental Risk Factors, and Seasonal Patterns of Dengue Virus Infection in Nigeria: A Systematic Review and Meta-analysis (2014–2024)

**DOI:** 10.64898/2026.02.28.26346557

**Authors:** Justin Onyebuchi Nwofe, Sunday Alapetei Gbeyedobo, Monday Tarshi, Queensly Onah, Praise Danson, Aisha Bisallah Aburke, Ogechukwu Nwakaego Onyebuchi, Adamu Ishaku Akyala

## Abstract

**Background:** Dengue virus (DENV) is an increasingly recognized cause of febrile illness in sub-Saharan Africa, yet its epidemiology in Nigeria remains incompletely characterized due to fragmented surveillance and diagnostic variability. We conducted a systematic review and meta-analysis to estimate marker-specific seroprevalence and to evaluate geographic variation, seasonal patterns, and environmental risk factors associated with DENV infection in Nigeria between 2014 and 2024.

**Methods:** Following PRISMA guidelines, we searched PubMed, Scopus, Web of Science, EMBASE, Google Scholar, and African Index Medicus for studies reporting laboratory-confirmed dengue infection in Nigeria. Random-effects meta-analysis was used to estimate pooled prevalence and pooled odds ratios (ORs) with 95% confidence intervals (CIs). Between-study heterogeneity was quantified using the I² statistic. Subgroup analyses explored variation by diagnostic marker and geographic region.

**Results:** Thirty-three studies met inclusion criteria. The pooled random-effects IgM seroprevalence was 20.9% (95% CI: 13.0–30.1; I² = 98.6%), while pooled IgG seroprevalence was 19.9% (95% CI: 11.6–29.7; I² = 97.7%). NS1 antigen positivity was 8.9% (95% CI: 2.2–19.4), and PCR-confirmed infection was 7.0% (95% CI: 1.2–16.2; I² = 25.8%). Significant differences were observed across diagnostic markers (p = 0.0002). Regional subgroup analysis demonstrated statistically significant geographic variation for both IgM (p = 0.0179) and IgG (p = 0.0030), with highest pooled prevalence observed in the Southeast and lowest in the Southsouth region. Environmental and behavioral exposures were strongly associated with seropositivity, including proximity to refuse dumpsites (OR = 9.39, 95% CI: 7.44–11.84), non-use of mosquito nets (OR = 8.70, 95% CI: 5.73–13.21), malaria positivity (OR = 5.54, 95% CI: 3.84–7.98), and open household water storage (OR = 2.18, 95% CI: 1.65–2.89). All four DENV serotypes were identified across reporting studies. Transmission intensity increased during rainy seasons.

**Conclusions:** Dengue virus transmission in Nigeria is widespread and geographically heterogeneous, with evidence of both recent and cumulative exposure. Strong associations with modifiable environmental and household-level factors underscore the importance of integrated vector control and improved diagnostic capacity. Enhanced surveillance and climate-informed public health strategies are essential to mitigate future outbreak risk.

**Author Summary:** Dengue is a mosquito-borne viral infection that is increasing globally but remains underrecognized in much of sub-Saharan Africa. In Nigeria, many febrile illnesses are presumed to be malaria, which can obscure the contribution of dengue virus infection. In addition, limited surveillance systems and inconsistent diagnostic testing have made it difficult to determine the true extent of dengue transmission. To address this gap, we conducted a systematic review and meta-analysis of studies published between 2014 and 2024 to evaluate patterns of dengue exposure, geographic variation, and environmental risk factors in Nigeria.

We found evidence of both recent infection (IgM antibodies) and past exposure (IgG antibodies) across multiple regions of the country. Transmission intensity varied geographically, with higher levels observed in some regions, particularly in the Southeast, and lower levels in the Southsouth. Infection risk increased during the rainy season, consistent with enhanced mosquito breeding conditions. Living near refuse dumpsites, storing water in open containers, not using mosquito nets, and having malaria were all associated with higher odds of dengue infection, highlighting the importance of household and environmental conditions in shaping transmission risk. All four dengue virus serotypes were identified, indicating sustained viral circulation.

These findings demonstrate that dengue virus infection is widespread in Nigeria and influenced by modifiable environmental and behavioral factors. Improving diagnostic capacity, strengthening routine surveillance, and implementing targeted vector control strategies are essential to reduce transmission and improve outbreak preparedness.

## Introduction

Dengue virus (DENV), a mosquito-borne flavivirus, has emerged as a major global public health concern, with incidence increasing over recent decades in association with rapid urbanization, population growth, and climate change (1,2). The World Health Organization estimates that approximately 3.9 billion people in more than 128 countries are at risk of infection, with nearly 390 million infections occurring annually, of which about 96 million manifest clinically(3,4). Although historically concentrated in Southeast Asia, the Western Pacific, and the Americas, dengue is increasingly reported across Africa and the Eastern Mediterranean, reflecting its expanding geographic range (3).

Dengue is characterized by the co-circulation of four antigenically distinct serotypes (DENV-1 to DENV-4). Secondary infection with a heterologous serotype may increase the risk of severe disease through antibody-dependent enhancement (5–9). Urbanization, inadequate infrastructure, poor drainage systems, ineffective waste management, and widespread household water storage create favorable breeding habitats for Aedes mosquitoes, thereby facilitating sustained transmission (2,10–12). Increased human mobility and regional connectivity further enhance viral dissemination across ecological zones (13–16).

Despite growing global recognition, dengue epidemiology in sub-Saharan Africa remains incompletely defined. In Nigeria, febrile illnesses are frequently presumed to be malaria and/or typhoid because of overlapping clinical features, potentially obscuring the contribution of arboviral infections (17–20). Studies conducted across West Africa report wide variability in dengue seroprevalence, suggesting ongoing but underrecognized transmission (21–24). Limited diagnostic capacity, inconsistent surveillance systems, and heterogeneous reporting frameworks complicate accurate estimation of national disease burden.

Nigeria presents ecological and demographic conditions conducive to dengue transmission. Rapid urban expansion, high population density, seasonal rainfall variability, and widespread peri-domestic vector habitats create environments favorable for Aedes proliferation (25–27). Reported seroprevalence estimates vary substantially across Nigerian states, reflecting differences in diagnostic approaches (e.g., IgM, IgG, NS1 antigen detection, and molecular assays), study populations, and sampling periods (28–33).

Recent molecular investigations have documented circulation of multiple dengue serotypes, with evidence of geographic clustering and regional viral movement within West Africa (34–36). supporting the possibility of sustained endemic transmission.

Seasonality represents a critical but under-synthesized dimension of dengue epidemiology in Nigeria. Vector abundance typically increases during rainy months, amplifying opportunities for viral transmission. In addition, environmental and household-level factors including water storage practices, waste accumulation, and vector control behaviors may significantly influence infection risk. However, these determinants have not been quantitatively integrated at the national level.

Given these gaps, a comprehensive synthesis of available evidence is needed to clarify the burden and determinants of dengue virus infection in Nigeria. Systematic review and meta-analysis provide a structured approach to integrate heterogeneous data, generate marker-specific pooled prevalence estimates, quantify between-study heterogeneity, and evaluate environmental and demographic correlations of infection. Therefore, this study conducted a systematic review and meta-analysis of laboratory-confirmed dengue infections reported in Nigeria between 2014 and 2024 to: (1) estimate pooled seroprevalence across diagnostic markers; (2) evaluate demographic and environmental risk factors through pooled effect-size analysis; and (3) characterize geographic and seasonal patterns relevant to transmission dynamics. By consolidating available evidence, this study aims to guide targeted improvements in dengue surveillance and outbreak monitoring, improve differential diagnosis of febrile illness, and support targeted, evidence-based vector control strategies.

## Materials and Methods

### Study Design and Reporting Framework

This study was conducted as a systematic review and meta-analysis to synthesize evidence on the seroprevalence, transmission patterns, and risk factors associated with dengue virus (DENV) infection in Nigeria between January 2014 and August 2024. The review protocol was developed prior to study initiation and prospectively registered in the International Prospective Register of Systematic Reviews (PROSPERO; Registration No. CRD420261328923) and is publicly accessible at https://www.crd.york.ac.uk/PROSPERO/myprospero. No amendments were made to the protocol following registration. Reporting followed the Preferred Reporting Items for Systematic Reviews and Meta-Analyses (PRISMA) 2020 guidelines (37).

### Eligibility Criteria

Studies were eligible if they reported laboratory-confirmed DENV infection among human populations residing in Nigeria and provided sufficient data to compute prevalence estimates or effect sizes. Acceptable diagnostic methods included serological detection of IgM or IgG antibodies, NS1 antigen assays, plaque reduction neutralization tests (PRNT), or molecular confirmation using reverse transcription polymerase chain reaction (RT-PCR). Observational study designs, including cross-sectional, cohort, case-control, and surveillance-based investigations, were included.

Studies conducted outside Nigeria, vector-only investigations without human infection data, editorials, commentaries, reviews, conference abstracts without accessible full text, and studies lacking laboratory confirmation were excluded.

### Search Strategy and Study Selection

A comprehensive literature search was conducted in PubMed, Scopus, Web of Science, EMBASE, Google Scholar, and African Index Medicus to identify studies reporting laboratory-confirmed dengue virus infection in Nigeria. The search covered publications from January 2014 to August 2024 and was restricted to studies involving human populations. Search terms combined controlled vocabulary (e.g., MeSH terms) and free-text keywords related to “dengue,” “dengue virus,” “seroprevalence,” “epidemiology,” and “Nigeria,” using Boolean operators (AND/OR) and database-specific filters where applicable. The complete search strategies for each database are provided in Supporting Information.

All retrieved records were imported into Covidence for duplicate removal and systematic screening. Two independent reviewers screened titles and abstracts against predefined eligibility criteria, followed by full-text assessment of potentially relevant articles. Disagreements were resolved through discussion and, when necessary, consultation with a third reviewer.

### Data Extraction

Data was extracted using a standardized form within Covidence. The primary outcome of interest was laboratory-confirmed dengue virus infection, defined by detection of IgM antibodies (recent infection), IgG antibodies (past exposure), NS1 antigen (acute infection), reverse transcription polymerase chain reaction (RT-PCR), or plaque reduction neutralization test (PRNT), as reported in individual studies. For each diagnostic marker, the number of positive cases and total sample size were extracted to permit calculation of marker-specific prevalence estimates. Where multiple diagnostic markers were reported within a study, data were extracted separately for each marker.

Secondary outcomes included environmental and behavioral risk factors associated with dengue seropositivity. For studies reporting exposure data, 2×2 contingency table information (exposed cases, exposed total, unexposed cases, unexposed total) was extracted to enable pooled odds ratio estimation. When studies reported multiple measures of association, raw frequency data were prioritized to ensure consistent effect-size computation across studies.

All compatible prevalence estimates reported within the defined study period were considered. Where studies reported results for multiple subgroups (e.g., geographic region, season, or demographic category), overall study-level estimates were preferentially extracted to maintain comparability across studies unless subgroup analysis was explicitly required.

Additional variables extracted included author, year of publication, geographic region, study design, study setting (hospital-based or community-based), sample size, population characteristics (e.g., age distribution), diagnostic method, serotype distribution where available, and reported environmental or household-level exposures. Information on funding sources was not recorded.

Data extraction was performed independently by two reviewers to ensure accuracy and completeness. Discrepancies were resolved through discussion. Where information was unclear or incomplete, study reports were reviewed in detail to identify extractable data; no imputation of missing prevalence or exposure data was performed. Studies lacking sufficient data to compute effect estimates were excluded from quantitative synthesis but described narratively where appropriate.

### Risk of Bias Assessment

Methodological quality was assessed using the Joanna Briggs Institute (JBI) Critical Appraisal Checklist for Studies Reporting Prevalence Data. Each study was evaluated across domains including sampling strategy, measurement validity, and statistical analysis. Discrepancies were resolved through consensus.

### Statistical Analysis

All statistical analyses were conducted using R software (version 4.5.2) with the *meta* package. Studies were grouped for synthesis according to diagnostic marker (IgM, IgG, NS1, PCR, PRNT) and separately for each risk factor where extractable 2×2 data were available. Only studies reporting sufficient marker-specific outcome data were included in the corresponding meta-analysis. Forest plots were generated to display individual study estimates and pooled effects. Funnel plots were constructed to assess small-study effects where ≥10 studies were available. Formal assessment of certainty of evidence using the Grading of Recommendations

Assessment, Development and Evaluation (GRADE) framework was not performed, as the included studies were predominantly observational prevalence studies and the review aimed to synthesize epidemiologic estimates rather than evaluate intervention effects.

### Pooled Prevalence Estimation

Marker-specific pooled prevalence estimates were calculated separately for IgM, IgG, NS1 antigen, and PCR-confirmed infection using random-effects meta-analysis. Variance-stabilizing transformations (Freeman–Tukey double arcsine) were applied prior to pooling of proportions. Random-effects models were estimated using the inverse variance method with Hartung–Knapp adjustment for confidence intervals.

Between-study heterogeneity was assessed using Cochran’s Q statistic and quantified using the I² statistic. Forest plots were generated for visualization.

### Subgroup Analysis

Subgroup analyses were conducted to explore potential sources of heterogeneity related to geographic regions and diagnostic methods. Statistical differences between subgroups were evaluated using χ² tests for subgroup interaction.

### Risk-Factor Meta-Analysis

For environmental and behavioral exposures, pooled odds ratios (ORs) with 95% confidence intervals were calculated using random-effects meta-analysis of 2×2 contingency tables. Models were estimated using the inverse variance method with Hartung–Knapp adjustment.

Heterogeneity was assessed using I² statistics. Sensitivity analyses were conducted using leave-one-out procedures to assess the influence of individual studies on pooled effect estimates.

### Publication Bias

Publication bias was assessed through visual inspection of funnel plots and Egger’s regression test when at least ten studies were available for a given marker.

### Serotype Distribution

Serotype data were summarized descriptively using aggregated counts across reporting studies due to limited availability of comparable molecular data.

## Results

### Study Selection

The database search yielded 1,115 records. After removal of duplicates, 817 records remained for screening. Title and abstract screening resulted in 71 articles undergoing full-text assessment. Thirty-three studies met the inclusion criteria and were included in the meta-analysis (Fig 1).

**Figure 4.1.**
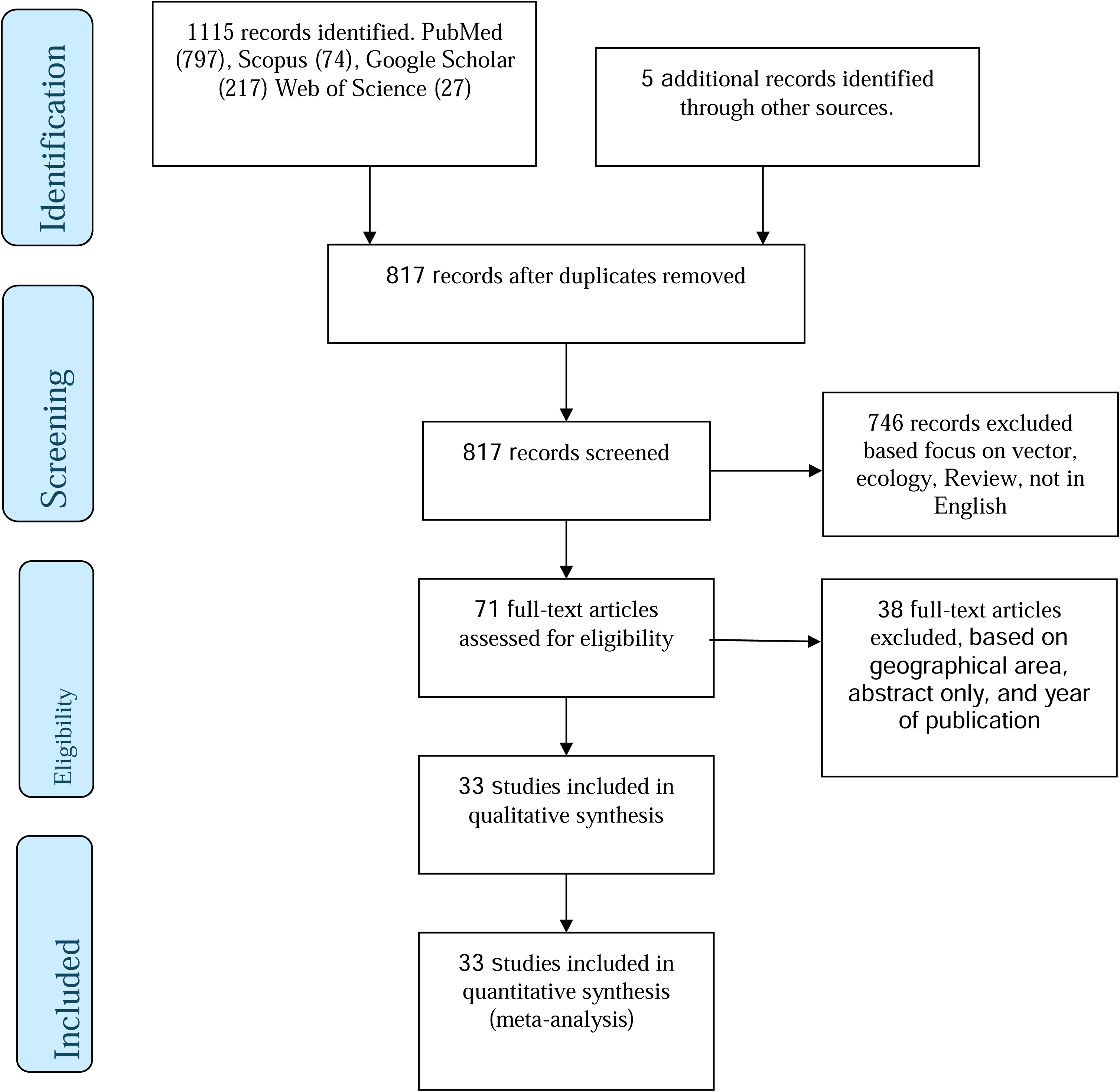
PRISMA flow diagram of search strategy for inclusion of published studies.

### Study Characteristics

The included studies were published between 2014 and 2024 and comprised predominantly cross-sectional designs conducted in hospital-based and community-based settings. Study populations included both febrile and non-febrile individuals across diverse ecological zones of Nigeria. Diagnostic methods included enzyme-linked immunosorbent assays (ELISA) for IgM and IgG antibodies, rapid diagnostic tests, NS1 antigen detection assays, plaque reduction neutralization tests (PRNT), and reverse transcription polymerase chain reaction (RT-PCR). Detailed characteristics of included studies are summarized in Table 1.

**Table 1:**
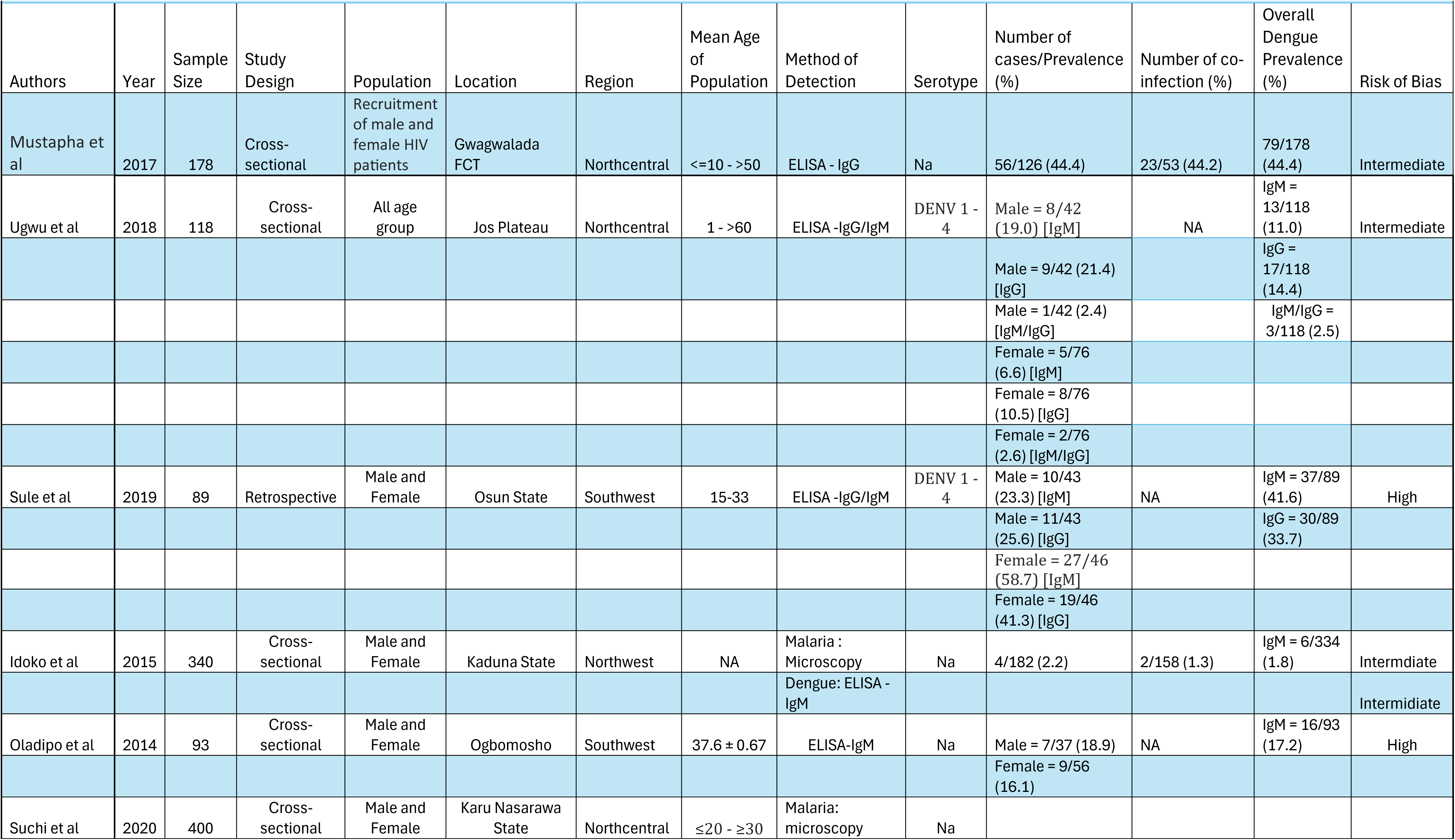
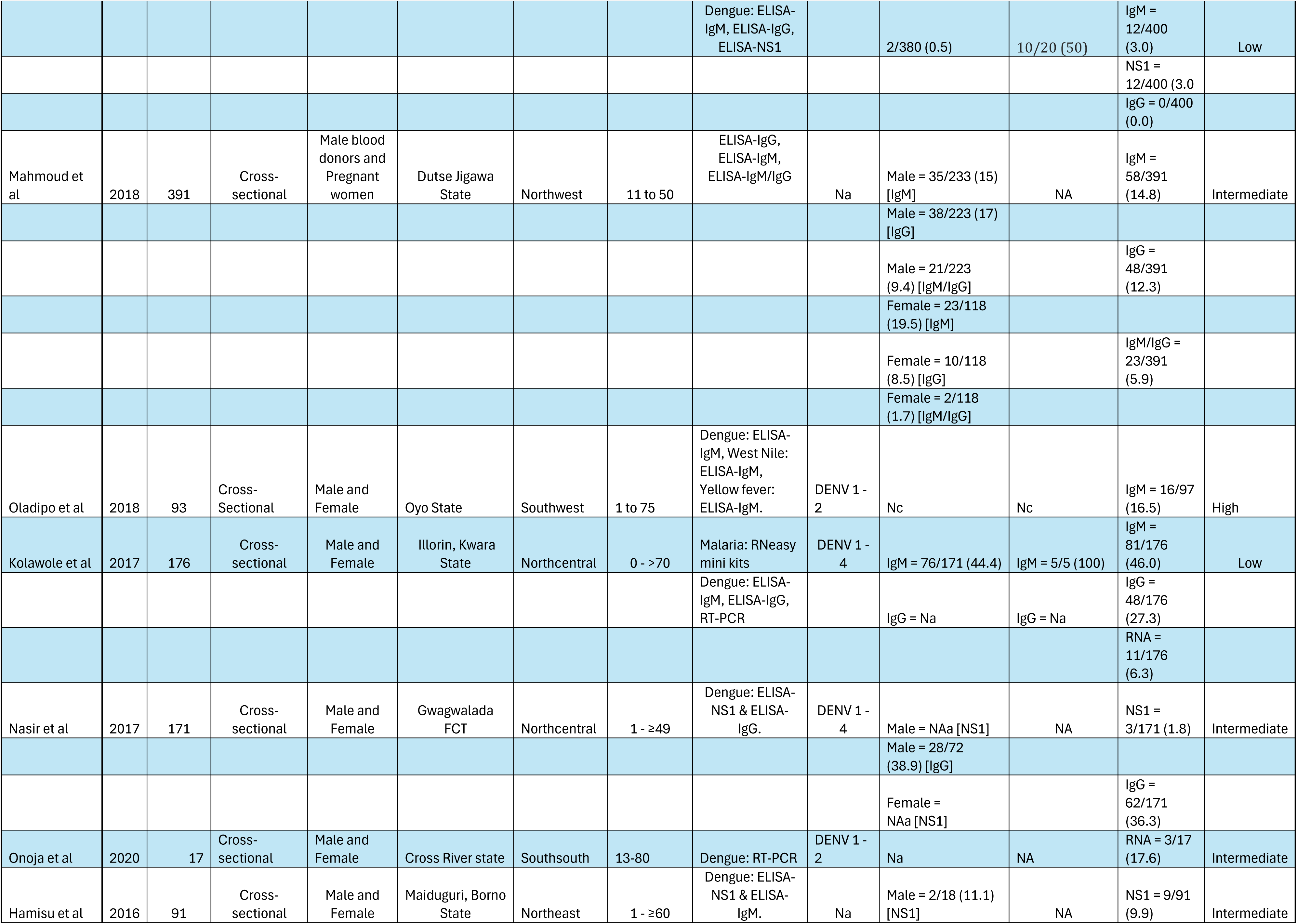
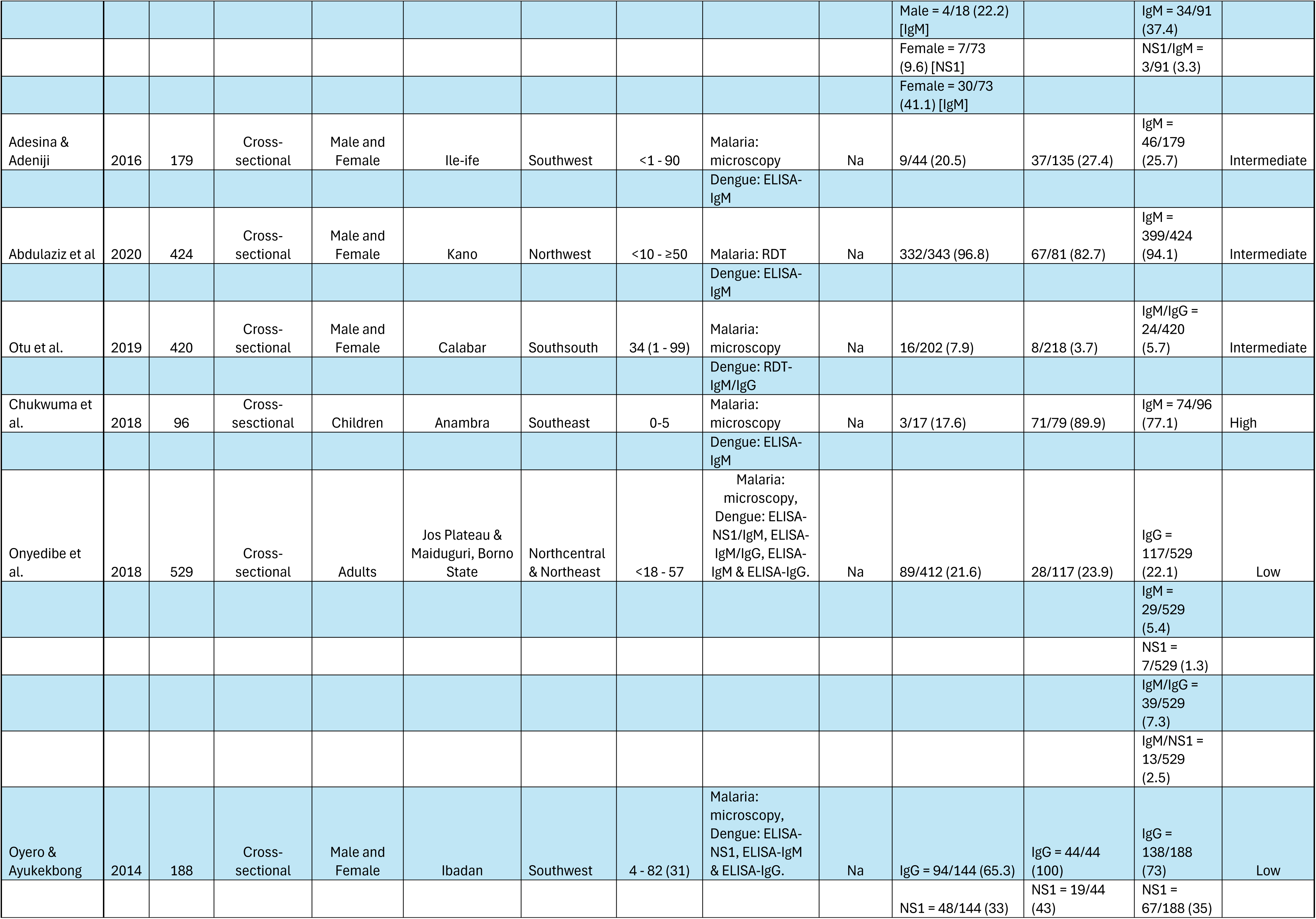
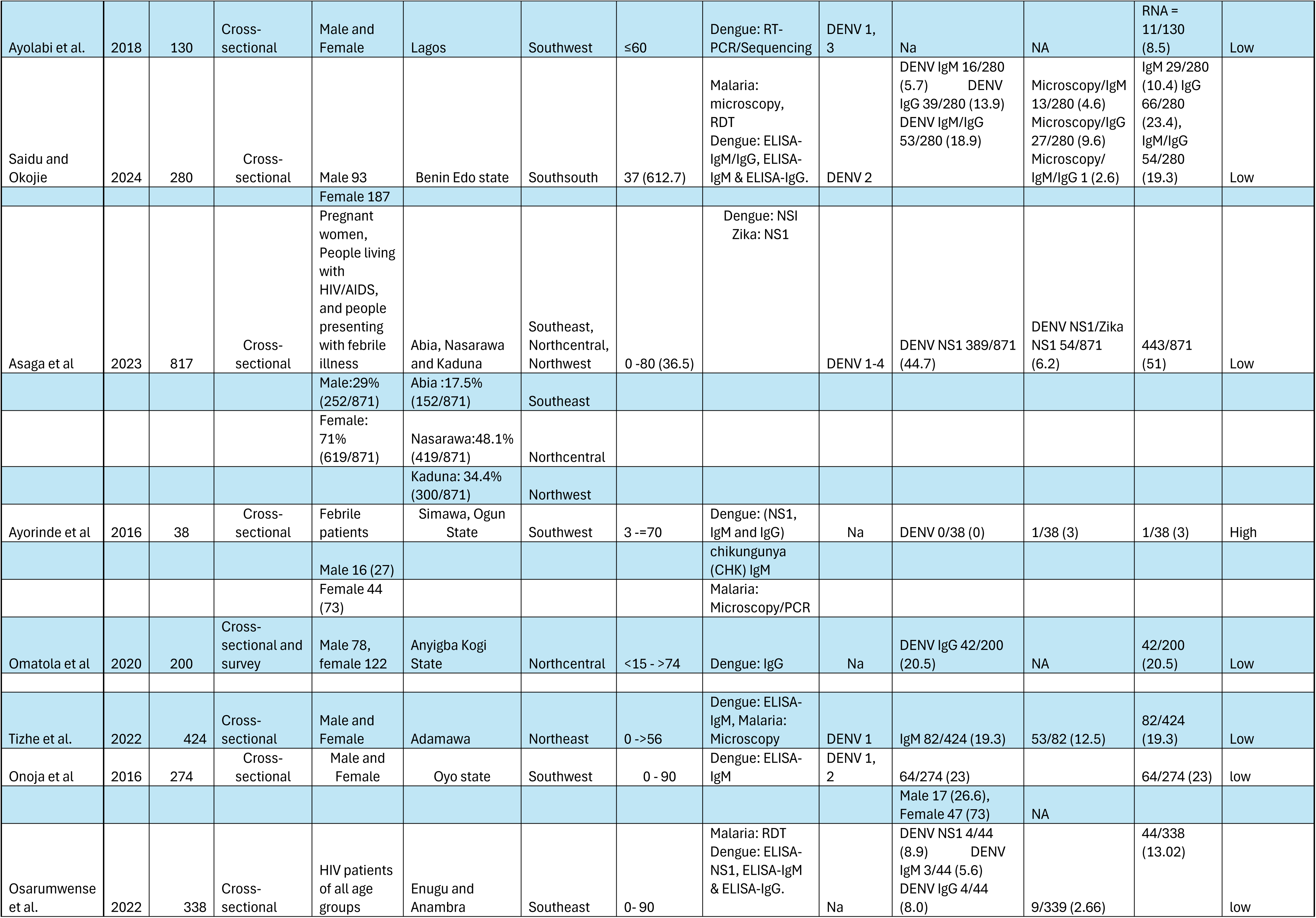
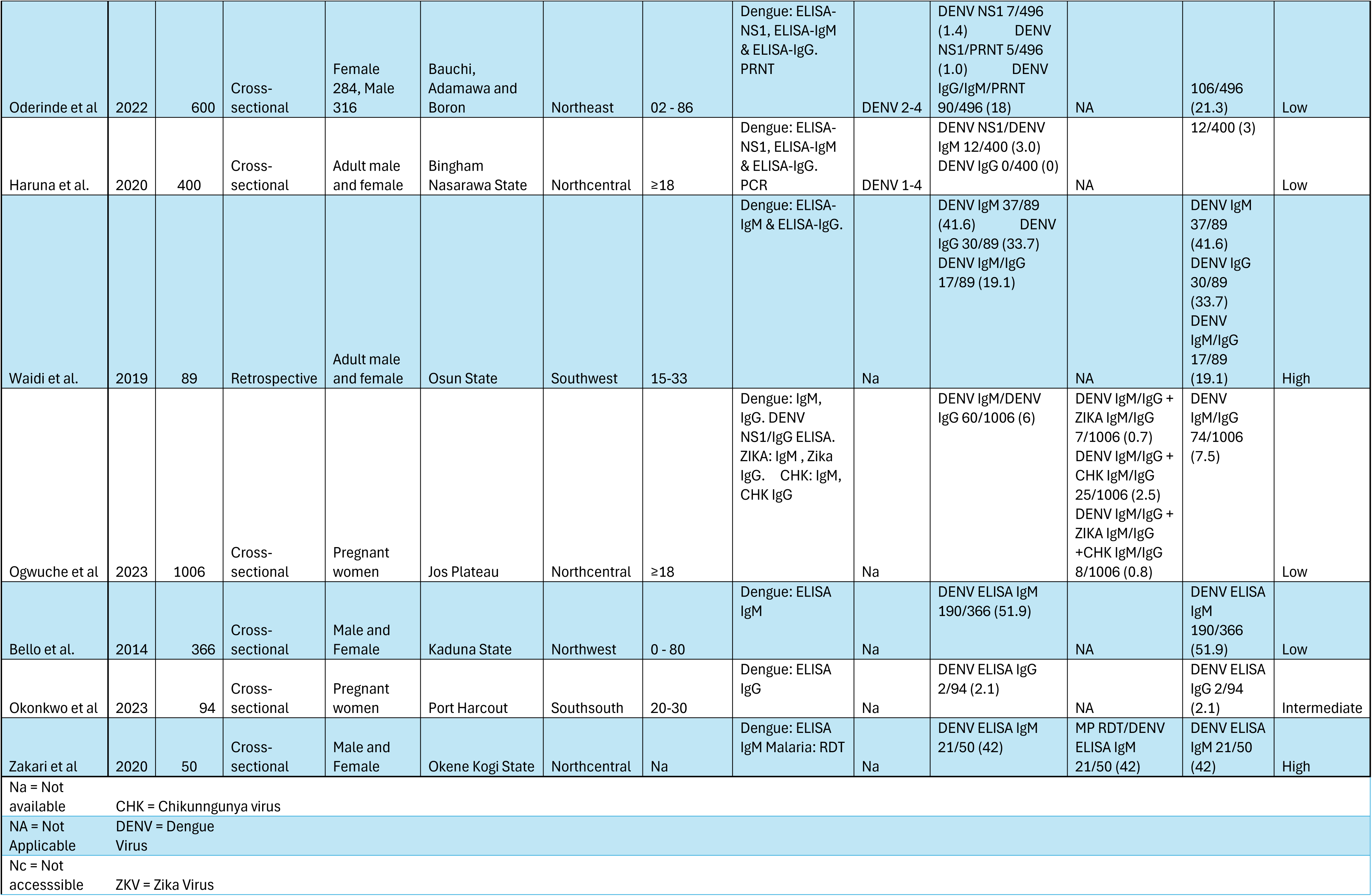
Study Characteristics included in the Systematic Review.

### Marker-Specific Pooled Prevalence IgM Seroprevalence

Twenty-six studies comprising 7,318 participants and 1,453 IgM-positive cases were included in the IgM meta-analysis. The pooled random-effects IgM seroprevalence was 20.9% (95% CI: 13.0–30.1). Substantial heterogeneity was observed (I² = 98.6%, τ² = 0.0653, Q = 1789.70, p < 0.001), supporting use of a random-effects model. Table 2.

**Table 2:** Risk Factors for Dengue Seropositivity.

| **Exposure** | **k (studies)** | **Pooled OR (Random)** | **95% CI** | **I<sup>2</sup> (%)** | **p-value** |
| --- | --- | --- | --- | --- | --- |
| Refuse dumpsite | 33 | 9.39 | 7.44–11.84 | 0 | <0.001 |
| Open water storage | 23 | 2.18 | 1.65–2.89 | 0 | <0.001 |
| No mosquito net use | 23 | 8.70 | 5.73–13.21 | 32 | <0.001 |
| Malaria positivity | 22 | 5.54 | 3.84–7.98 | 23.8 | <0.001 |

### IgG Seroprevalence

Seventeen studies including 4,861 participants and 831 IgG-positive cases were analyzed. The pooled random-effects IgG seroprevalence was 19.9% (95% CI: 11.6–29.7), with considerable heterogeneity (I² = 97.7%, τ² = 0.0474, Q = 698.36, p < 0.001).

### NS1 Antigen Positivity

Eleven studies (3,078 participants; 389 events) reported NS1 antigen detection. The pooled random-effects NS1 prevalence was 8.9% (95% CI: 2.2–19.4). Heterogeneity was high (I² = 98.5%, τ² = 0.0500, Q = 687.64, p < 0.001).

### PCR-Confirmed Infection

Three studies comprising 323 participants and 25 PCR-confirmed cases were included. The pooled random-effects prevalence of PCR-confirmed dengue infection was 7.0% (95% CI: 1.2–16.2), with low to moderate heterogeneity (I² = 25.8%, Q = 2.70, p = 0.26).

One study reported PRNT-confirmed neutralizing antibodies with a prevalence of 18.2%. Due to the availability of a single study, pooled meta-analysis was not performed for PRNT.

Subgroup comparison demonstrated significant differences in pooled prevalence across diagnostic methods (χ² = 21.55, df = 4, p = 0.0002), indicating variation in estimated burden depending on the laboratory marker used.

### Regional Variation in Seroprevalence

Regional subgroup analysis demonstrated statistically significant geographic variation in both IgM (χ² = 13.66, p = 0.0179) and IgG (χ² = 17.92, p = 0.0030) seroprevalence. For IgM, pooled random-effects prevalence ranged from 8% in the Southsouth to 37% in the Southeast. For IgG, pooled prevalence ranged from 9% in the Southsouth to 36% in the Southeast. These findings indicate consistent geographic gradients in both recent and past dengue virus exposure across Nigeria.

### Risk Factors for Dengue Seropositivity

Random-effects meta-analysis of environmental and behavioral exposures demonstrated strong and statistically significant associations.

Living near refuse dumpsites was associated with markedly increased odds of dengue seropositivity (OR = 9.39, 95% CI: 7.44–11.84; I² = 0%, p < 0.001). Leave-one-out sensitivity analysis confirmed stability of this association.

Open household water storage was associated with increased odds of infection (OR = 2.18, 95% CI: 1.65–2.89; I² = 0%, p < 0.001). Individuals not using mosquito nets had significantly higher odds of seropositivity compared with those using nets (OR = 8.70, 95% CI: 5.73–13.21; I² = 32.0%, p < 0.001). Malaria-positive individuals demonstrated higher odds of dengue seropositivity compared with malaria-negative individuals (OR = 5.54, 95% CI: 3.84–7.98; I² = 23.8%, p < 0.001). Table 3.

**Table 3:** Marker-Specific Pooled Prevalence.

| Marker | k | Events | Total | Random Prevalence (%) | 95% CI | I<sup>2</sup> (%) |
| --- | --- | --- | --- | --- | --- | --- |
| IgM | 26 | 1453 | 7318 | 20.9 | 13.0–30.1 | 98.6 |
| IgG | 17 | 831 | 4861 | 19.9 | 11.6–29.7 | 97.7 |
| NS1 | 11 | 389 | 3078 | 8.9 | 2.2–19.4 | 98.5 |
| PCR | 3 | 25 | 323 | 7.0 | 1.2–16.2 | 25.8 |

Across these exposures, heterogeneity ranged from absent to moderate, indicating consistent associations across studies.

### Serotype Distribution

Among studies reporting serotype-specific data, all four dengue virus serotypes (DENV-1 to DENV-4) were identified. Aggregated counts indicated that DENV-1 accounted for approximately 35% of serotyped cases, followed by DENV-3 (30%), DENV-2 (25%), and DENV-4 (10%) indicating sustained viral circulation and potential risk for secondary infections. However, serotype reporting was limited to a subset of studies, and proportions should be interpreted cautiously.

### Seasonal Patterns

Several studies reported higher prevalence estimates during rainy seasons compared with dry periods, consistent with increased vector breeding and seasonal amplification of transmission intensity.

### Publication Bias

Funnel plot inspection and Egger’s regression test for IgM seroprevalence did not demonstrate statistically significant small-study effects (p = 0.19).

## Discussion

This systematic review and meta-analysis provide quantitative evidence that dengue virus infection is widely distributed across Nigeria, with pooled seroprevalence estimates indicating substantial recent and cumulative exposure. The pooled random-effects estimates demonstrated substantial serologic evidence of DENV exposure in Nigeria. IgM seroprevalence was 20.9% (95% CI: 13.0–30.1), indicating recent transmission, while IgG seroprevalence was 19.9% (95% CI: 11.6–29.7), reflecting cumulative exposure. NS1 antigen positivity (8.9%) and PCR-confirmed infection (7.0%) further support ongoing viral circulation. The high heterogeneity observed across serologic markers (I² > 97%) likely reflects true ecological and epidemiologic variation across Nigeria’s diverse transmission settings. The observed prevalence levels are consistent with reports from other West African settings where dengue transmission has been historically underrecognized but increasingly documented (38). Similar patterns of geographic heterogeneity and diagnostic variability have been described across sub-Saharan Africa, reflecting differences in ecological suitability, urbanization intensity, and surveillance capacity. The detection of all four dengue serotypes further aligns with regional molecular investigations demonstrating sustained circulation and potential for endemic transmission. Together, these findings support growing evidence that dengue is not sporadic in West Africa but represents an established and underdiagnosed contributor to febrile illness within malaria-endemic settings (39).

### Spatial Distribution and Transmission Potential

Regional subgroup analysis demonstrated statistically significant geographic variation for both IgM and IgG. For IgM, pooled prevalence ranged from 8% in the Southsouth to 37% in the Southeast, while IgG ranged from 9% to 36% across the same regions. The consistent geographic gradient observed across both markers suggests sustained transmission in certain zones rather than isolated outbreaks. This spatial heterogeneity is consistent with patterns observed in other African settings where micro-ecological differences generate localized transmission hotspots. (40,41). Environmental suitability, urban density, and waste management infrastructure are likely to contribute to these regional differences. Nigeria’s rapid urban expansion, particularly in densely populated areas with inadequate drainage systems, may create ideal breeding habitats for Aedes mosquitoes. Human mobility and inter-state travel may further facilitate viral dissemination across ecological zones.(33,42,43).

### Environmental and Household Determinants of Infection

One of the most striking findings of this study was the magnitude and consistency of associations between modifiable environmental exposures and dengue seropositivity. Living near refuse dumpsites was associated with nearly ninefold higher odds of infection (OR = 9.39, 95% CI: 7.44–11.84), with no detectable heterogeneity across studies. Non-use of mosquito nets was associated with an 8.7-fold increase in odds (OR = 8.70, 95% CI: 5.73–13.21), while malaria positivity was associated with a 5.5-fold increase (OR = 5.54, 95% CI: 3.84–7.98). Open household water storage also significantly increased infection risk (OR = 2.18, 95% CI: 1.65–2.89).

The absence or low levels of heterogeneity across these environmental exposures strengthen the inference that these factors represent stable and reproducible determinants of dengue transmission within Nigerian settings. Refuse accumulation and water storage practices likely facilitate the proliferation of container-breeding Aedes mosquitoes (44,45). The strong association with malaria positivity underscores the diagnostic and ecological overlap between febrile illnesses in endemic areas and reinforces the importance of improved differential diagnosis.

### Seasonal Influence on Transmission

Seasonal amplification of dengue during rainy periods was consistently reported. Rainfall increases the availability of breeding habitats, while temperature influences viral replication within mosquito vectors. These findings align with established climate-driven transmission models and suggest that Nigeria’s bimodal rainfall patterns may create cyclical periods of elevated transmission risk. As climate variability intensifies, these seasonal peaks may become more pronounced or prolonged (46,47).

### Serotype Circulation and Implications for Disease Severity

All four DENV serotypes (DENV-1 to DENV-4) were identified among reporting studies, with DENV-1 and DENV-3 representing the largest proportions of typed samples. The co-circulation of multiple serotypes raises concern for antibody-dependent enhancement and the potential for severe disease during secondary infections. However, limited serotype reporting and inconsistent molecular surveillance highlight a need for expanded virological monitoring.

### Diagnostic Variability and Surveillance Gaps

Diagnostic methods varied substantially across studies, and subgroup analysis demonstrated significant differences in pooled prevalence across markers. Serologic assays showed higher heterogeneity compared with molecular detection, likely reflecting cross-reactivity and variation in assay performance. Standardization of diagnostic protocols and increased access to molecular testing are essential for improving data comparability and surveillance accuracy.

### Implications for Practice, Policy, and Future Research

The findings of this review have important implications for clinical practice and public health policy in Nigeria. The high pooled seroprevalence and evidence of seasonal amplification suggest that dengue represents a substantial and underrecognized contributor to febrile illness. Integration of dengue diagnostics into routine febrile illness evaluation, particularly during rainy seasons and in high-burden regions, may improve case detection and reduce misclassification as malaria. Identification of modifiable environmental risk factors including water storage practices and proximity to refuse sites supports the implementation of targeted, community-level vector control interventions.

At the policy level, the marked geographic heterogeneity observed underscores the need for region-specific surveillance strategies rather than uniform national approaches. Climate-informed monitoring systems may enhance early warning and outbreak preparedness. Strengthening laboratory infrastructure and standardizing diagnostic algorithms will be essential to improving comparability of epidemiologic data across states.

Future research should prioritize longitudinal cohort studies to better characterize temporal transmission dynamics, expanded molecular surveillance to clarify serotype distribution and viral evolution, and integrated entomological–epidemiological investigations to quantify vector–human interactions. Standardized reporting frameworks and consistent use of validated diagnostic tools will be critical to reducing heterogeneity and improving evidence quality.

### Limitations of the Included Evidence

This review provides the first comprehensive quantitative synthesis of marker-specific dengue seroprevalence and environmental risk factors in Nigeria. However, there are several limitations of the underlying evidence and should be considered when interpreting these findings. First, substantial heterogeneity was observed across studies, reflecting variability in study populations, geographic coverage, diagnostic approaches, and sampling periods. The predominance of cross-sectional and hospital-based designs may limit generalizability to the broader population, particularly in rural or underserved regions. Diagnostic variability including differences in sensitivity and specificity across IgM, IgG, NS1, and RT-PCR assays may have influenced prevalence estimates and contributed to between-study heterogeneity. Additionally, limited availability of standardized serotype data constrained more detailed analysis of serotype-specific transmission patterns. These factors underscore the need for harmonized surveillance and standardized diagnostic protocols in future investigations.

### Limitations of the Review Process

This review also has methodological limitations. Although comprehensive searches were conducted across multiple databases and grey literature sources, restriction to English-language publications may have introduced language bias. Despite efforts to identify unpublished and regionally indexed studies, some relevant data may have been missed. The reliance on published summary data limited the ability to perform individual participant–level analyses or adjust for confounding variables beyond reported aggregate measures. Furthermore, while publication bias was assessed using funnel plots and Egger’s regression test, such methods have limited power when the number of studies is small or heterogeneity is high. Formal certainty-of-evidence assessment using GRADE methodology was not performed, given the predominance of observational prevalence studies and the descriptive epidemiologic focus of the review.

## Conclusion

This systematic review and meta-analysis demonstrate that dengue virus transmission is widespread across Nigeria, with substantial evidence of both recent (IgM) and cumulative (IgG) exposure across multiple regions. The detection of all four dengue serotypes, combined with geographically consistent serologic gradients, indicates sustained viral circulation rather than isolated outbreaks. Significant regional variation in seroprevalence underscores the influence of ecological and environmental context on transmission dynamics. Strong and consistent associations with modifiable exposures including proximity to refuse dumpsites, non-use of mosquito nets, open household water storage, and malaria positivity highlight the central role of household and peri-domestic environments in shaping infection risk. Seasonal amplification during rainy periods further supports the climatic sensitivity of transmission.

Collectively, these findings suggest that dengue represents an underrecognized but established contributor to febrile illness in Nigeria. Diagnostic overlaps with malaria, limited molecular surveillance, and heterogeneity in testing approaches continue to obscure the true burden of infection. Strengthening laboratory capacity, integrating dengue diagnostics into routine febrile illness management, and expanding structured surveillance systems are critical to improving case detection and outbreak preparedness.

Future research should prioritize longitudinal surveillance, expanded molecular and serotype characterization, and standardized diagnostic protocols to enhance comparability across regions. Targeted, climate-informed vector control interventions particularly in rapidly urbanizing settings will be essential to mitigating transmission risk.

By synthesizing a decade of evidence, this study provides a robust epidemiologic foundation to inform policy, strengthen surveillance, and guide strategic public health responses to dengue in Nigeria.

## Acknowledgments

The authors would like to thank the Global Health and Infectious Diseases Control Institute of Nasarawa State University Keffi for their support and feedback on the design of this systematic review.

## Author Contributions

Conceptualization: Justin Onyebuchi Nwofe

Data curation: Justin Onyebuchi Nwofe, Sunday Gbeyedobo, Monday Tarshi, Queensly Onah, Praise Danson, Aisha Bisallah Aburke, Ogechukwu Nwakaego Onyebuchi

Formal analysis: Justin Onyebuchi Nwofe

Investigation: Justin Onyebuchi Nwofe, Sunday Gbeyedobo, Monday Tarshi, Queensly Onah, Praise Danson, Aisha Bisallah Aburke, Ogechukwu Nwakaego Onyebuchi

Methodology: Justin Onyebuchi Nwofe Software: Justin Onyebuchi Nwofe

Validation: Sunday Gbeyedobo, Monday Tarshi, Queensly Onah, Praise Danson, Aisha Bisallah Aburke, Ogechukwu Nwakaego Onyebuchi

Supervision: Adamu Ishaku Akyala Visualization: Justin Onyebuchi Nwofe

Writing – original draft: Justin Onyebuchi Nwofe, Sunday Gbeyedobo, Monday Tarshi, Queensly Onah, Praise Danson, Aisha Bisallah Aburke, Ogechukwu Nwakaego Onyebuchi

Writing – review & editing: Justin Onyebuchi Nwofe, Adamu Ishaku Akyala

## Data Availability Statement

The extracted study-level dataset used for meta-analysis, 2×2 contingency tables for risk-factor analyses, and analytic code (R script) are provided as Supporting Information files (S2–S5 Files). The review protocol is publicly available via PROSPERO (CRD420261328923). No individual participant data was used in this review.

---

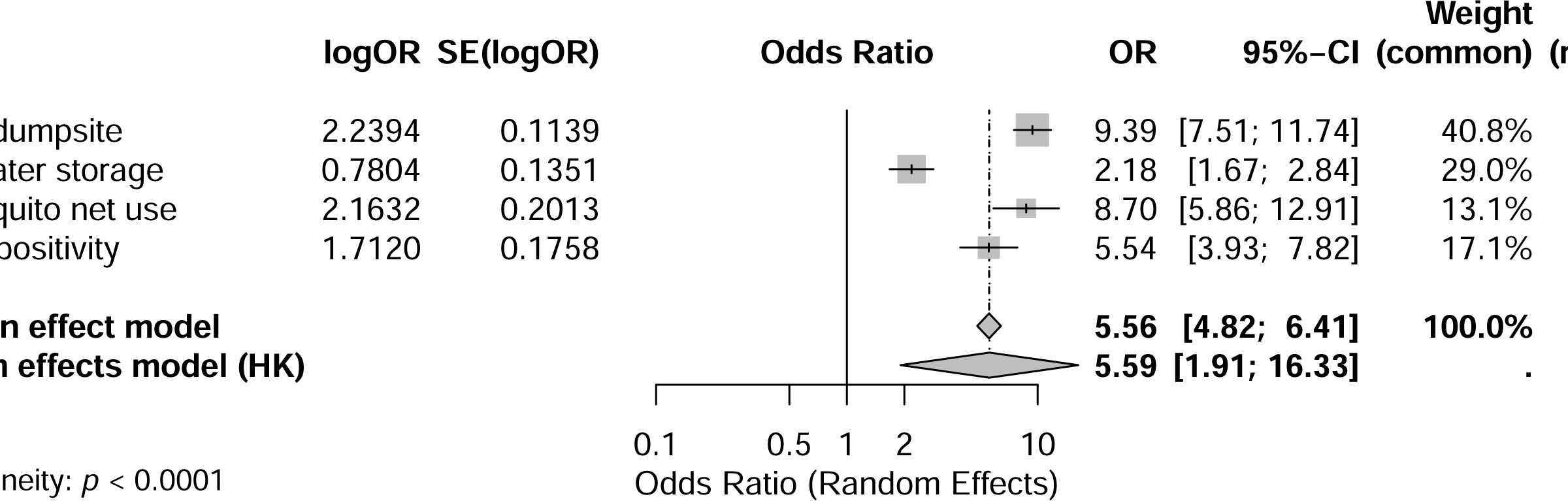

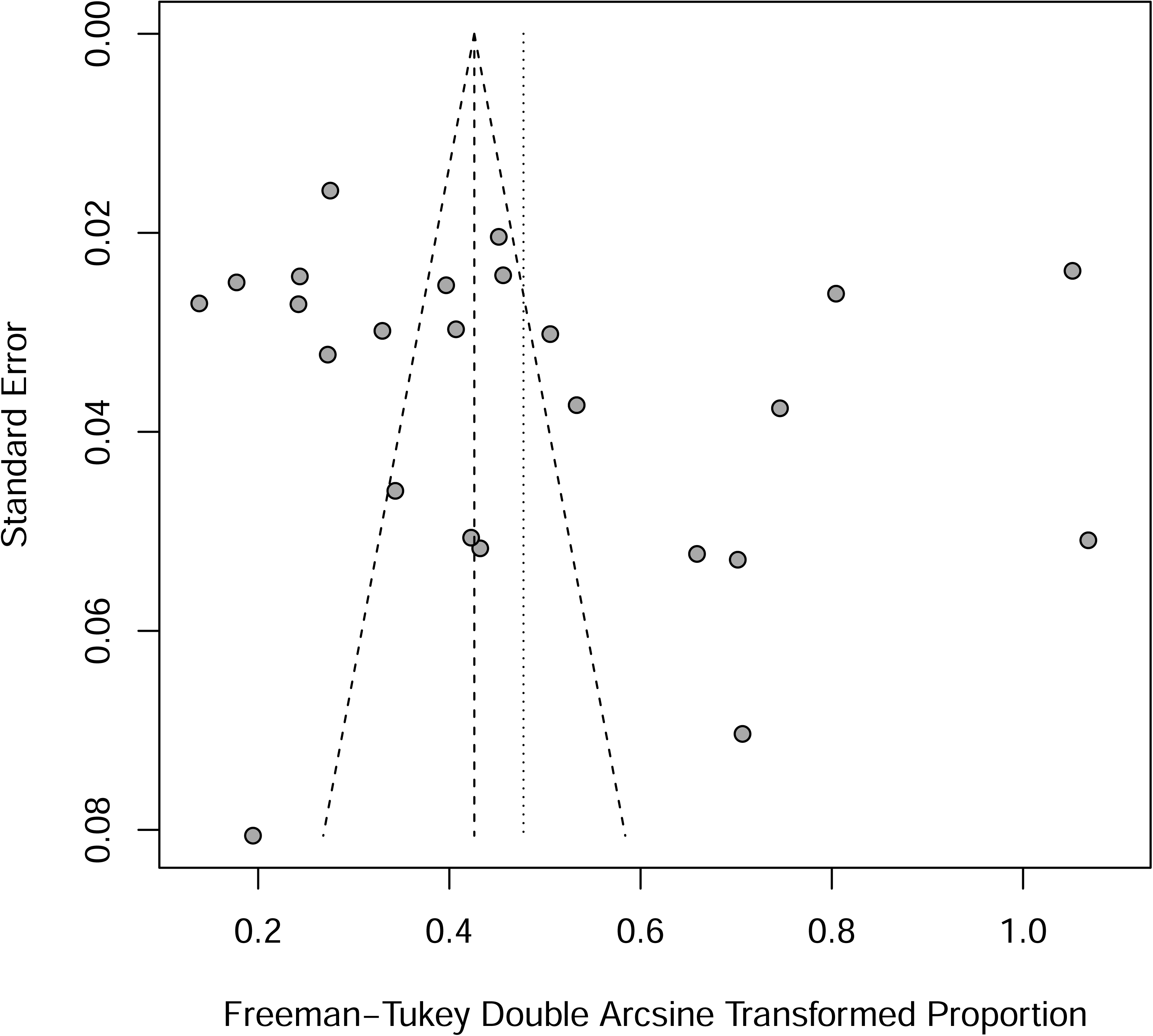

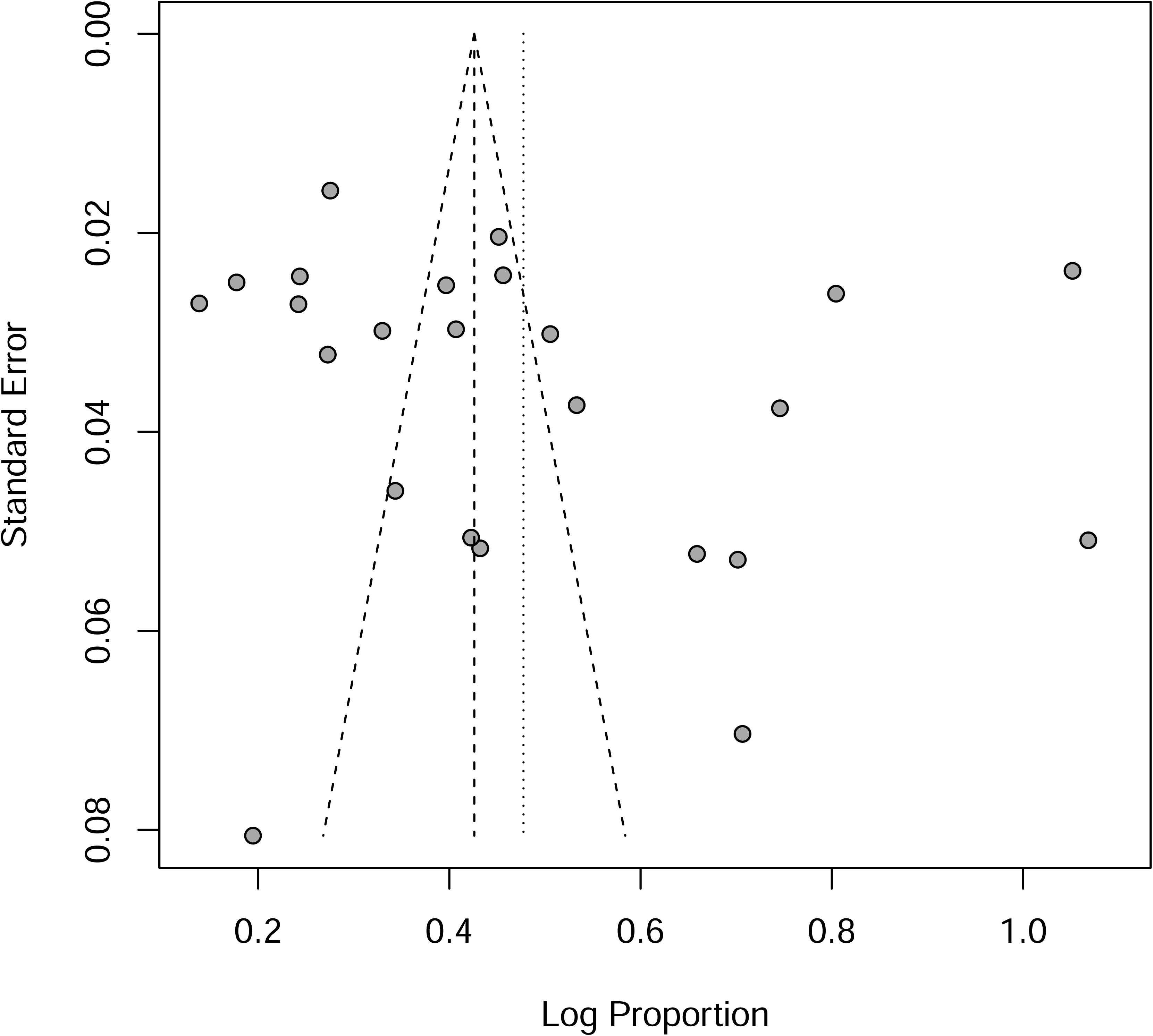

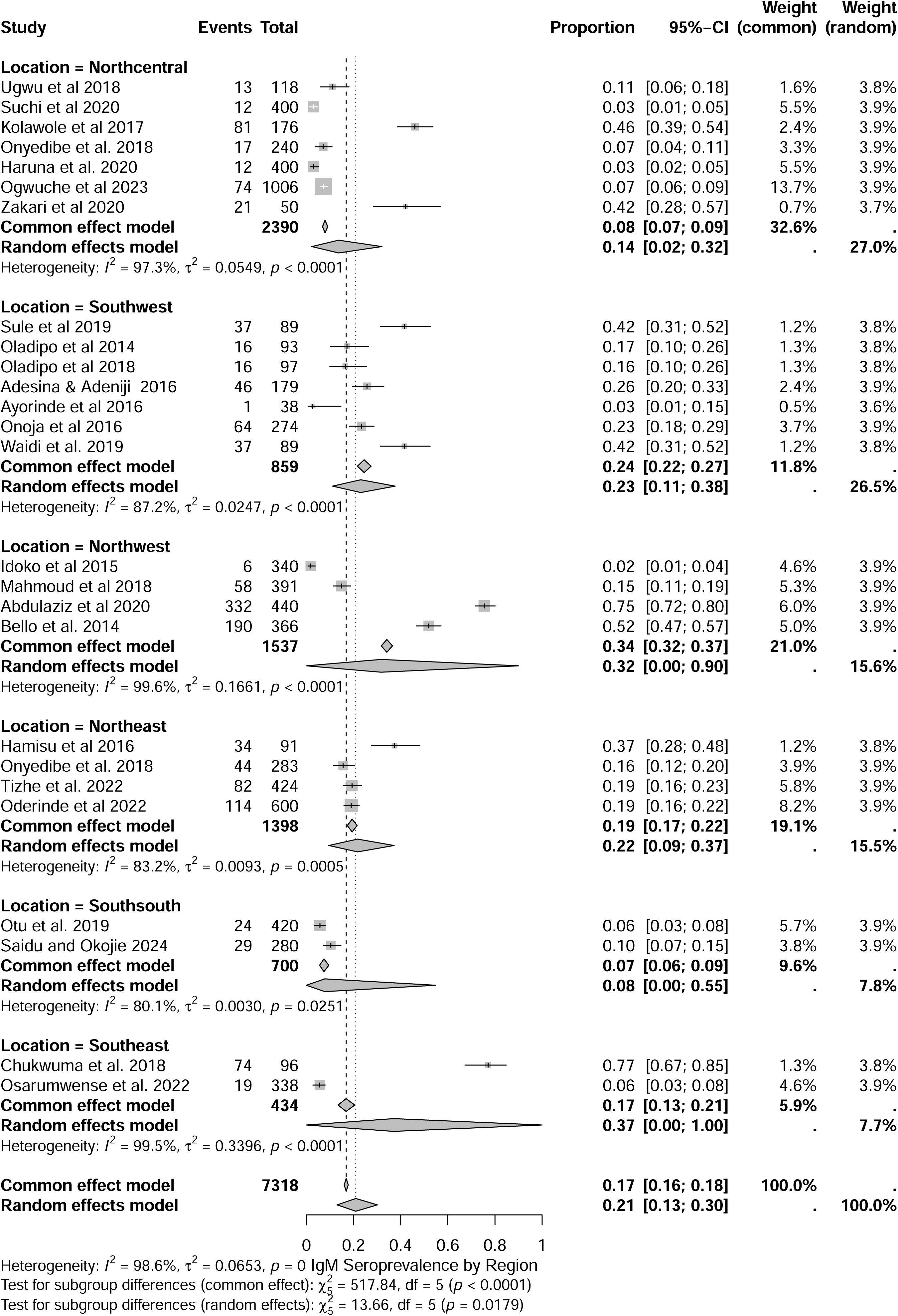

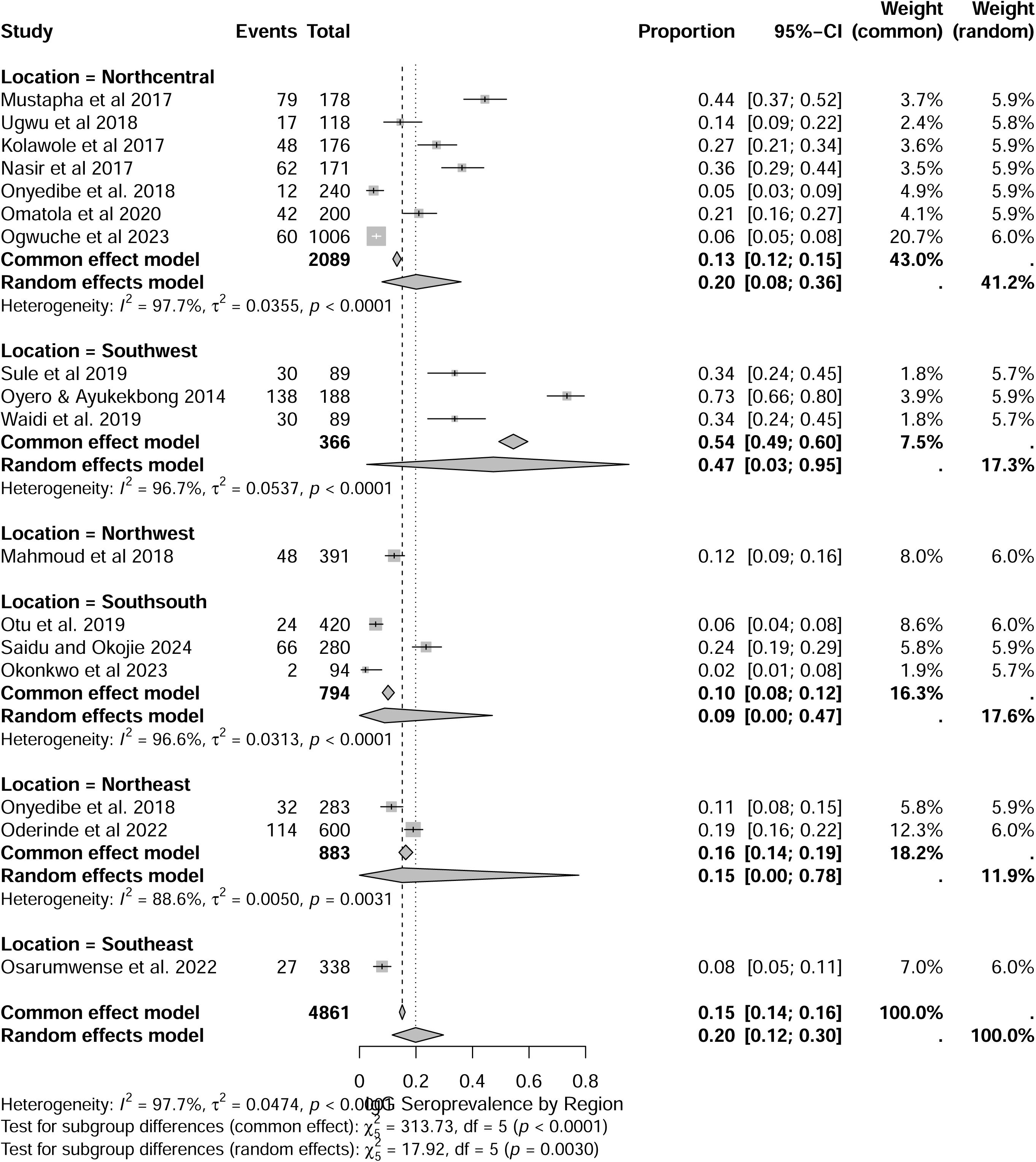

